# Evaluation of Public Awareness, Knowledge and Attitudes towards Basic Life Support among Non-Medical, Adult population in Muscat City, Oman: Cross-Sectional Study

**DOI:** 10.1101/2020.05.16.20104323

**Authors:** Aisha Aldhakhri, Gu Can

## Abstract

**OBJECTIVES:** This study investigated the level of public awareness, knowledge and attitudes towards BLS among non-medical adult Omanis in Muscat City, Oman and explored the association between knowledge and attitude and the socio-demographic characteristics.

**DESIGN:** Cross sectional design was used. Convenience sampling method in Muscat city from February to March 2020.

**Setting:** The study setting was based in Muscat City, Oman.

**Participants:** 426 Adults, non-medical Omanis; able to read and write Arabic Language; were included between February and March 2020.

**METHODS:** Adopted, validated, online Arabic questionnaire was used, including four parts of 37 questions on socio-demographic information, awareness, knowledge and attitude. The results were presented in tables with descriptions.

**FINDINGS:** Majority of participants were in age groups, 28-37 years (50.0%) and 18-27 years (35.7%), were female (57.0%); married (58.5%), single (39.9%); had secondary (31.5%), diploma (27.2%), bachelor’s (32.4%) education levels; and worked in government (28.6%), private sector (25.4%). Significantly, large proportion of them (62.0%) were aware about BLS. However, knowledge total scores weren’t normally distributed (0.917, P<0.001), with minimum, maximum, median and mean knowledge total score of 0 out of 15, 13 out of 15, 3 out of 15, 3.6 ± 1.9 respectively. Also, attitude total scores weren’t normally distributed (0.976, P<0.001), with minimum, maximum, median and mean attitude total scores of 30 out of 65, 65 out of 65, 55 out of 65, 54.5 ± 5.5 respectively.

**CONCLUSIONS:** The awareness of nonmedical adults toward Basic Life Support was substantial, whereas, knowledge level toward BLS was very low despite of its differences with respect to socio-demographic characteristics. In contrast, all participants had positive attitude toward BLS. Therefore, this study is advocating the need for mandatory training in Basic Life Support for all non- medics in the country as well as incorporating Basic Life Support teaching in school curriculum.

## Introduction

Basic life support (BLS) is a level of medical care that is offered to victims of life-threatening illnesses or injuries until they can be given full medical care at a hospital.^1^ Considering that life-threatening illnesses or injuries may occur at places where there are no medical practitioners or health service providers, it is imperative that laypeople are equipped with all the necessary knowledge and skills to provide BLS. Nevertheless, there is a tendency of people to make assumptions that BLS ought to be performed by qualified medical personnel and this has resulted in some deaths that could have been avoided. Therefore, it is necessary to assess the general public on their knowledge, and awareness about BLS; and their attitude toward BLS so that proper interventions could be performed as appropriate.^2^ In the Oman population, the annual incidence of OHCA was reported in 2017, and it stated that the annual incidence ranged between 516–780 cases.^3^ Furthermore, the outcomes and coronary angiography findings of patients following OHCA in Oman indicated that 13% of patients survived and were discharged, although three survivors suffered from permanent hypoxic brain damage.^3^ Further, the Sultan Qaboos University Hospital in Oman observed that virtually every cardiac arrest patient admitted was brought to the hospital by friends or family relatives without being given cardiopulmonary resuscitation (CPR) first aid. Therefore, the foregoing situation in Oman may be attributed to lack of adequate knowledge, awareness, and skills of the general public in performing BLS.

## Methods

### Participants and Design

A total of 426 non-medical Omani participants were included; who aged 18 or more; willing to participate in the study; and able to speak and read Arabic or English languages. Particularly, data collection took virtually one month from 28^th^ February, 2020 until 28^th^ March, 2020. However, cross sectional design was used and subjects were collected using non-probability convenience sampling technique. Therefore, the interested and eligible participants were approached every day during the study period from 9 am up until 6 pm in Sultan Qaboos University (SQU) public areas, waiting areas in Sultan Qaboos University Hospital (SQUH) and other public places like shopping malls, sports facilities, parks, and other entertainment areas. A website-based questionnaire tool was easily accessed by the subjects.

### Setting

The study setting was based in Muscat City, the Capital and largest City of the Sultanate of Oman where the majority of the population resides approximately 1,475,000 according to 2017 statistics. Therefore, those people were selected in the study to represent the study population, which can reflect positively on the sampling representativeness and generalizability. Thus, this gave more chance in attaining the required sample and increased variation of the measured variables.

### Instrument

A specific, validated Arabic questionnaire was developed, which consisted of four sections. The first part which contains socio-demographic information, was designed by the researcher. This part consisted of five questions that tapped information regarding participants’ age, gender, marital status, education, and occupation. The second part consisted of four questions asking about participant’s awareness of BLS. The third part was about participant’s knowledge of BLS that has 15 multiple choice questions. This validated part of the questionnaire was adopted from the Author, Dr.Maha Al-Mohaissen, who had developed the original questionnaire and used it in her study, Knowledge and Attitude Toward Basic Life Support Among Health Students at a Saudi Women’s University; a cross-sectional study, which was conducted in Suadi Arabia in 2016.^4^ The questionnaire with its key answers conformed to the latest American Heart Association (AHA) BLS guidelines.The fourth part, Attitude toward BLS, was about participants’ attitude toward BLS which was Likert Rating Scale with five levels (1=Strongly disagree, 2=disagree, 3=uncertain, 4=agree, and 5=strongly agree). It was adopted from the author Dr. Anurag Patidar who used it in his study; Attitude of school students towards Basic Life Support in Punjab, India, 2014.^5^ The questionnaire was modified by consulting with five PhD experts from both the clinical and academic nursing departments, Oman. CVI result was good with an average validity score of 92%. Also, test retest reliability coefficient of the attitude scale was calculated as r=0.784. Both Forward-Backward translation with the validity of the accuracy of wording were done. The tool was tested on 20 eligible subjects prior to data collection. A website-based questionnaire tool was designed and easily accessed by the participants.

### Analysis

The SPSS program was used to analyze the data. A qualified statistician and data manager were employed to manage and analyze these data appropriately. Categorical variables were summarized as counts and percentages, whereas continuous variables, including knowledge and attitude, were summarized as means and corresponding standard deviations, or medians and interquartile range (IQR). Moreover, knowledge total scores and attitude total scores were tested for normality using the Shapiro-Wilk test and their distributions were compared between the categories of socio-demographic characteristics. Thus, the student t-test or analysis of variance (ANOVA) was performed to compare the distributions when data were normally distributed; otherwise the Mann Whitney U test or the Kruskal Wallis H test was used. Furthermore, the difference between proportions within a categorical variable was tested using the homogeneity test of proportions, while the differences in the distributions of knowledge and attitude between the categories of a socio-demographic characteristic were assessed using independent samples tests and, where necessary, the post hoc analysis was performed to detect the significance of the differences between paired comparisons, for which the Bonferroni correction was used to adjust the *P* values. All the statistical tests were two-tailed and the significance level was set at the 5%.

### Patient/Public Involvement

Patients and public were not invited to comment on the study design, outcomes or interpret the results or writing or editing the document.

## Results

### Sample characteristics

Majority of the subjects were in the age groups, 28-37 years (50.0%) and 18-27 years (35.7%). In addition, majority of the subjects were female (57.0%); married (58.5%) or single (39.9%); had secondary (31.5%), diploma (27.2%), or bachelor’s (32.4%) levels of education; and worked in government (28.6%) or private sector (25.4%) (**Table 1**).

**Table 1:**
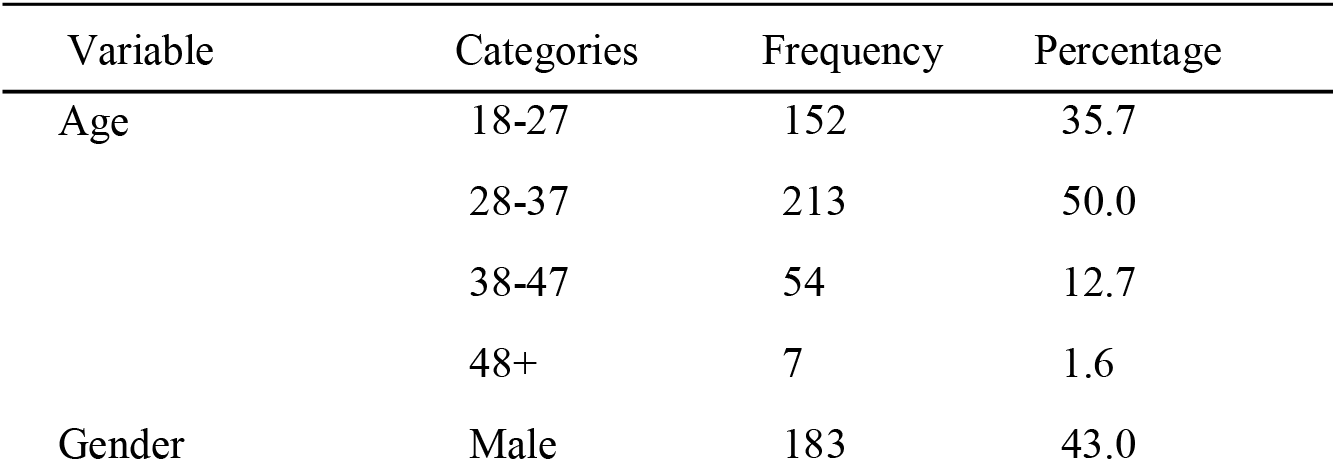
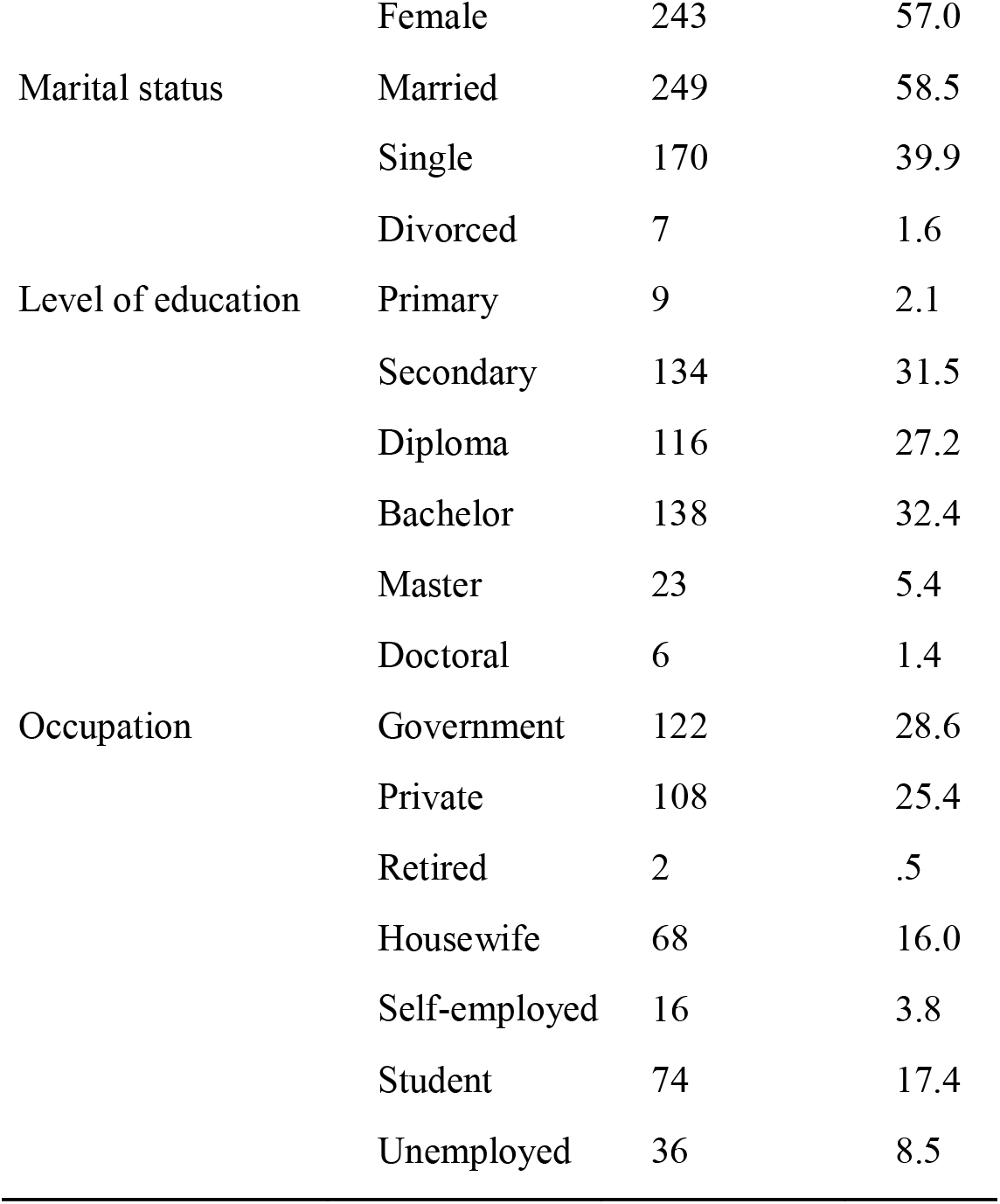
Sample Characteristics

### Participant’s awareness

Large proportion (62.0%) were already aware about cardio-pulmonary resuscitation (CPR), but had never received any CPR training (70.9%). Nevertheless, majority of those who indicated that they had received CPR training (29.1%) specified that they had received the training from Television-Internet-Media (33.1%), at a course given by the trainers of the Ministry of Health (21.0%), CPR education institutions (12.9%) and at school (12.1%). Moreover, those who indicated that they had not received CPR training expressed that they intend to attend CPR training in the future (84.8%). **(Table 2)**

**Table 2:**
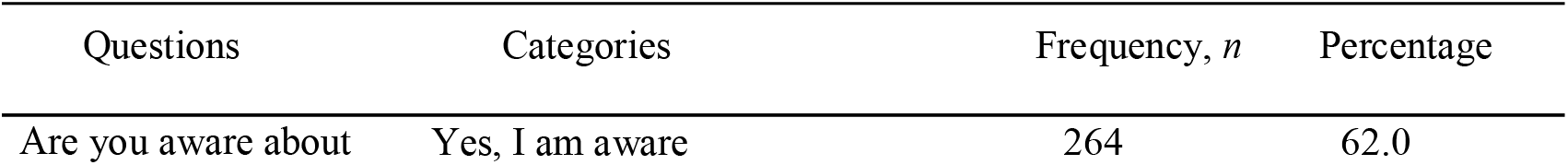
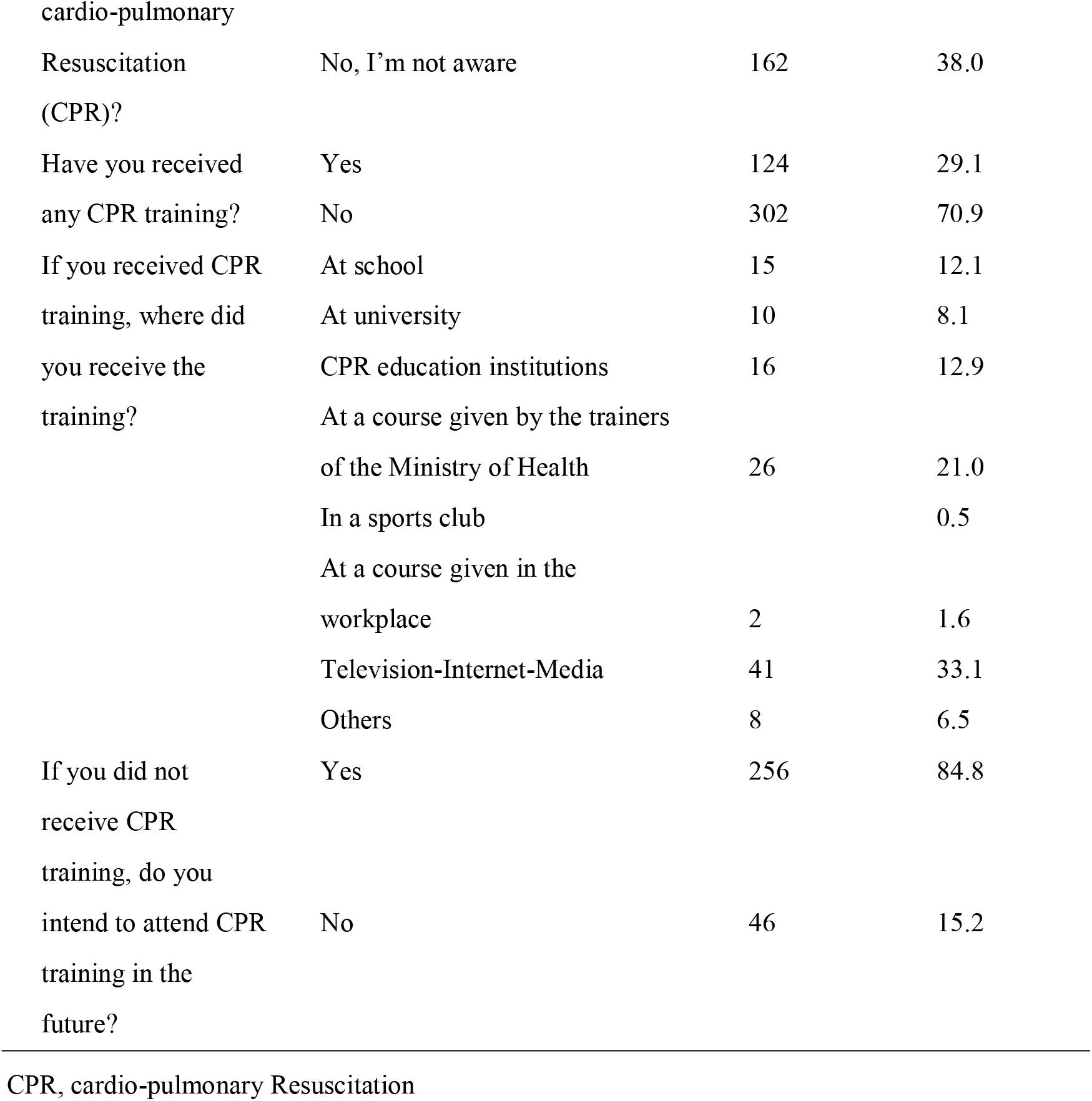
Public A wareness About BLS

### Participant’s knowledge

Generally, smaller proportions of the participants responded correctly on how to find out whether collapsed person was conscious [132 (31.0%)]; how to find out if a collapsed person had a pulse [162 (38.0%)]; and the location for chest compressions [139 (32.6%)]. However, larger proportions responded correctly on what to do next as an individual after confirming that the collapsed person was unconscious, not breathing and had no pulse [222 (52.1%)]; what number to call for emergency medical services [328 (77.0%)]; and what to do first upon witnessing an infant choking while playing with a toy but the infant was unable to cry or cough [241 (56.6%)] (**Table 3)**.

**Table 3:** Public knowledge about BLS

| Question | Frequency, *n* | Percentage |
| --- | --- | --- |
| What to do when you see a person collapsed on the road |  |  |
| (a) Correct response | 42 | 9.9 |
| (b) Incorrect response | 384 | 90.1 |
| How would you find out whether collapsed person was conscious? |  |  |
| (a) Correct response | 132 | 31.0 |
| (b) Incorrect response | 294 | 69.0 |
| How would you find out if a collapsed person had a pulse |  |  |
| (a) Correct response | 162 | 38.0 |
| (b) Incorrect response | 264 | 62.0 |
| If You confirm that the collapsed person is unconscious, not breathing and has no pulse. What would you do next? (Note: You are alone) |  |  |
| (a) Correct response | 222 | 52.1 |
| (b) Incorrect response | 204 | 47.9 |
| What number would you call for emergency medical services? |  |  |
| (a) Correct response | 328 | 77.0 |
| (b) Incorrect response | 98 | 23.0 |
| What is the location for chest compressions for adults? |  |  |
| (a) Correct response | 139 | 32.6 |
| (b) Incorrect response | 287 | 67.4 |
| What is the correct rate of chest compression for adults and children? |  |  |
| (a) Correct response | 48 | 11.3 |
| (b) Incorrect response | 378 | 88.7 |
| What is the correct depth of chest compression for adults |  |  |
| (a) Correct response | 33 | 7.7 |
| (b) Incorrect response | 393 | 92.3 |
| What is the correct ratio of Cardio-Pulmonary- Resuscitation (Compression: Ventilation Ratio) for an adult when there is a single rescuer? |  |  |
| (a) Correct response | 48 | 11.3 |
| (b) Incorrect response | 378 | 88.7 |

**Table 3** *Continued*
| Question | Frequency, *n* | Percentage |
| --- | --- | --- |
| What is the correct depth of chest compression for children and infants? |  |  |
| (a) Correct response | 8 | 1.9 |
| (b) Incorrect response | 418 | 98.1 |
| What is the location for chest compressions in an infant? |  |  |
| (a) Correct response | 40 | 9.4 |
| (b) Incorrect response | 386 | 90.6 |
| How do you give rescue breaths to infants? |  |  |
| (a) Correct response | 29 | 6.8 |
| (b) Incorrect response | 397 | 93.2 |
| If you and your friend are having meal and your friend suddenly starts expressing symptoms of choking, what should your first response be? |  |  |
| (a) Correct response | 17 | 4.0 |
| (b) Incorrect response | 409 | 96.0 |
| If you are witnessing an adult unresponsive victim who has been submerged in fresh water and just removed and just removed from it He has spontaneous breathing, but he is unresponsive. What is the first step? |  |  |
| (a) Correct response | 28 | 6.6 |
| (b) Incorrect response | 398 | 93.4 |
| If you witness an infant who suddenly starts to choke while playing with a toy. You have confirmed that he is unable to cry or cough. What should your first response be? |  |  |
| (a) Correct response | 241 | 56.6 |
| (b) Incorrect response | 185 | 43.4 |

### Distribution of knowledge total scores

Knowledge total scores weren’t normally distributed (W (426) = 0.917, P<0.001), with minimum knowledge total score of 0 out of 15, maximum knowledge total score of 13 out of 15, median knowledge total score of 3 out of 15 (IQR: 2 - 4), and mean ± SD knowledge total score of 3.6 ± 1.9. Note that SD stands for Standard Deviation and IQR stands for Inter Quartile Range. In addition, majority of the participants scored 3 out of 15 (27.0%), followed by 4 out of 15 (21.4%), then 2 out of 15 (17.4%). Therefore, majority of the participants had low knowledge about BLS.

### Distribution of attitude total scores

Attitude total scores of the participants weren’t normally distributed *(W* (426) =0.976, *P*<0.001), with minimum attitude total score of 30 out of 65, maximum attitude total score of 65 out of 65, median attitude total score of 55 out of 65 (IQR: 51 - 58), and mean ± SD attitude total score of 54.5 ± 5.5. Also, item scores, for each attitude item were not normally distributed (*W* (426) = 0.976, P<0.001). Therefore, majority of the participants had positive attitude towards BLS (**Table 4)**.

### Distribution of knowledge and attitude across socio-demographic variables

Considering that both BLS knowledge and attitude total scores were either not normally distributed, or measured on a smaller number of subjects in the respective categories of the socio-demographic variables, non-parametric tests were used to assess the significance of any difference in distributions across the foregoing socio-demographic variable categories. Therefore, the independent samples Kruskal Wallis H test was used to compare the distributions among more than two categories, whereas the Mann Whitney U test was used to compare the distributions between two categories. The results indicated that, although the participants were generally poor in knowledge about BLS, male participants were more knowledgeable than female participants. In addition, those with Doctoral level of education were more knowledgeable than those who had other levels of education. Besides, those with occupation in Government and Private sector were more knowledgeable than housewives and students (**Tables 5**). However, the level of attitude was the same across the categories of the socio-demographic variables **(Table 6)**. Besides, there was a significant weak positive linear relationship between the knowledge total scores and the attitude total scores (Pearson correlation: r = 0.116, P value = 0.017).

**Table 5:**
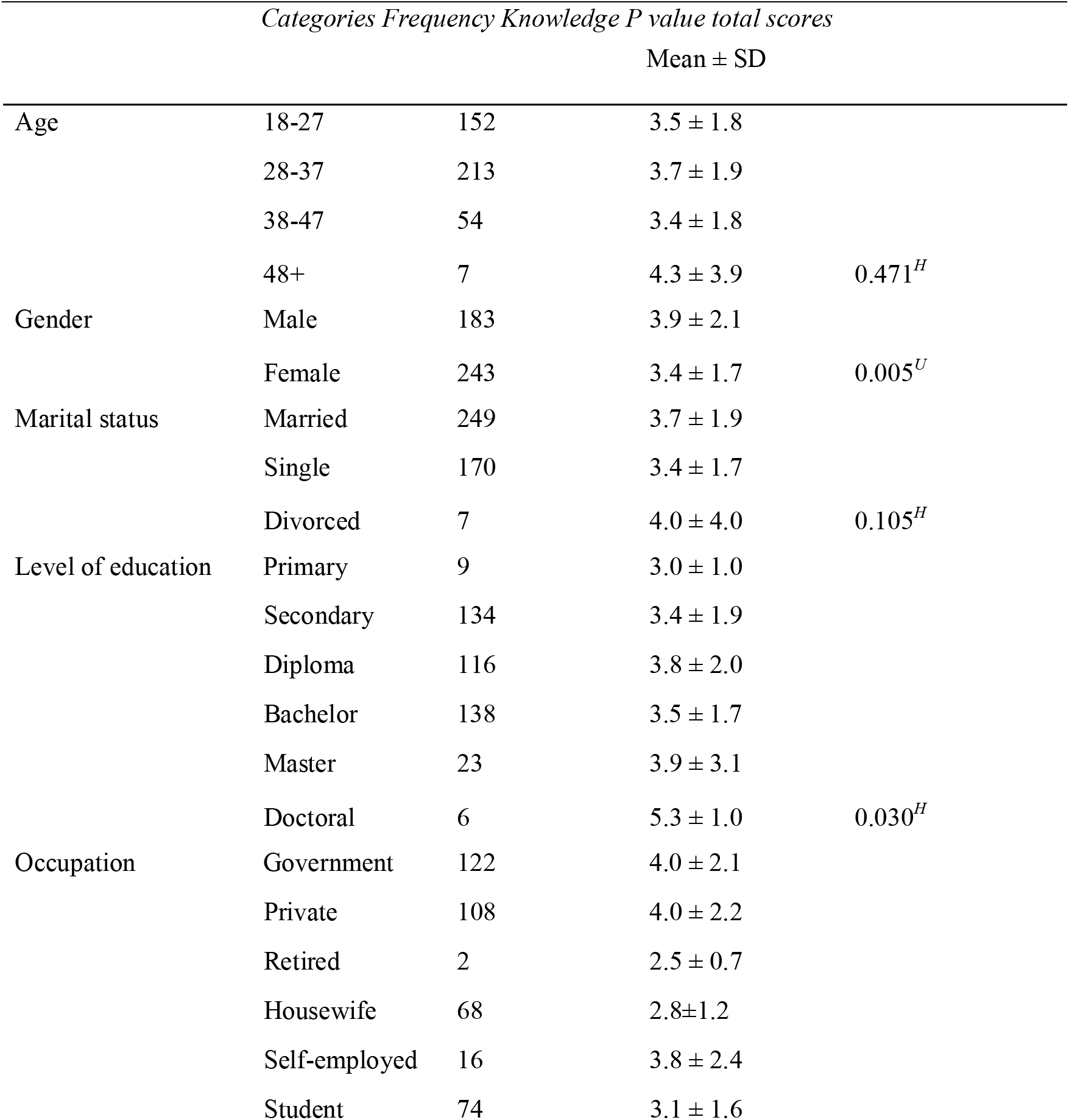
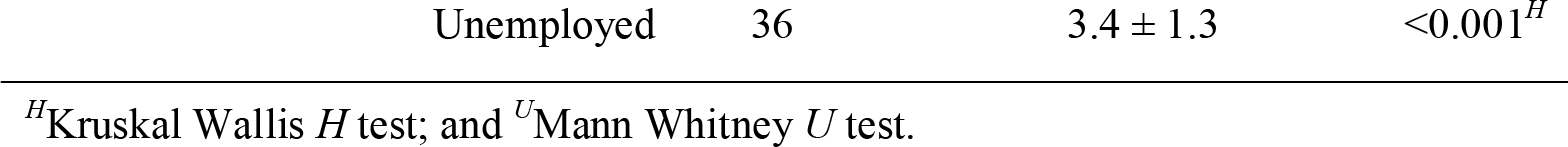
Distribution of BLS Knowledge Across the Socio-Demographic Variables

**Table 6.**
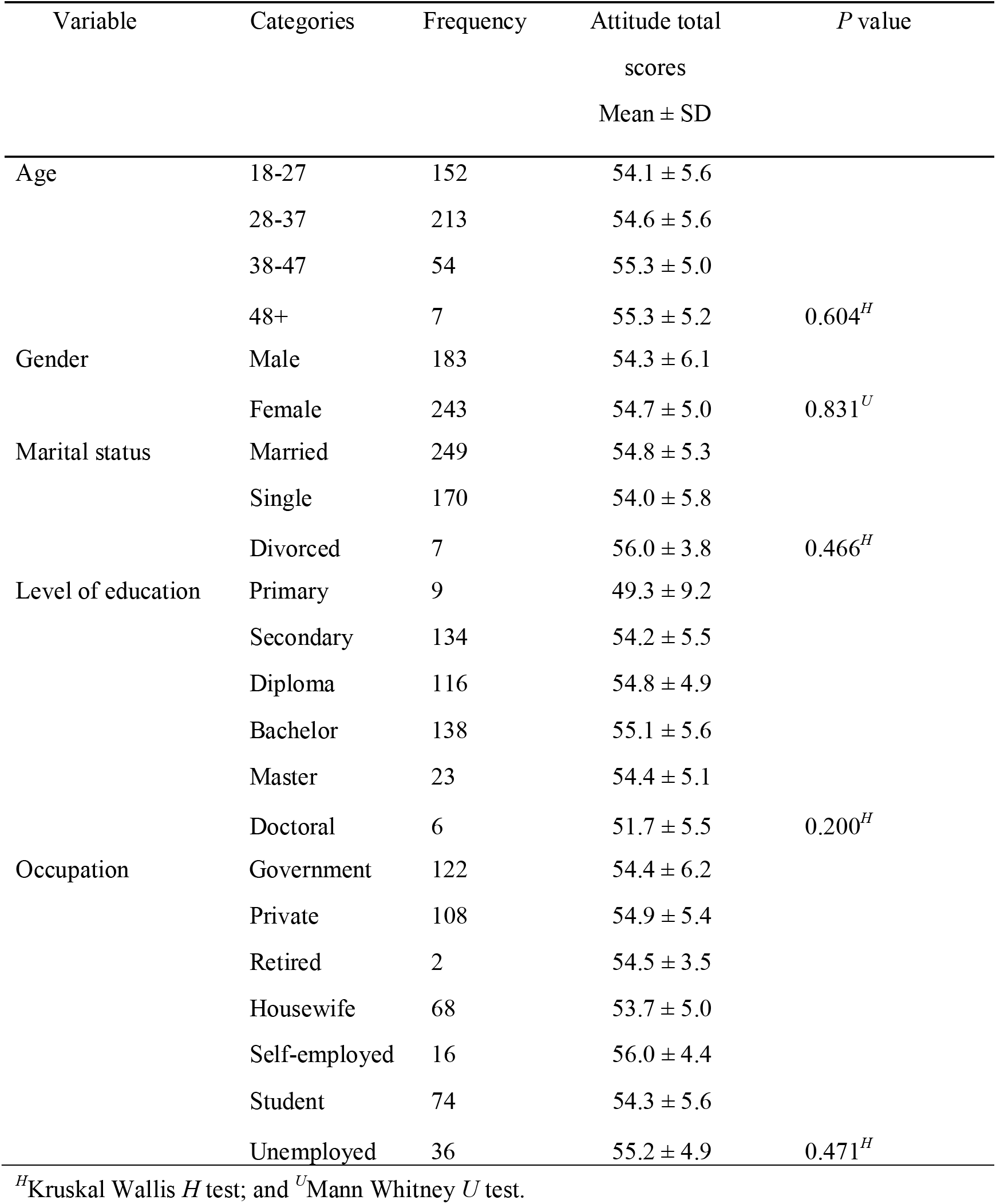
Distribution of BLS attitude across the socio-demographic variables

## Discussion

### Public awareness

Although a cross-sectional survey, which was carried out in 5 cities in the Sultanate of Oman, found that there is low effort in making the general public in Oman aware of the importance of CPR, which was manifested by the 60% of lay people surveyed, who indicated that they did not know how to perform CPR ^6^ this study found that a large proportion of the participants (62.0%) were already aware about cardio-pulmonary resuscitation (CPR).

Despite having awareness about CPR, 70.9% of this proportion had indicated that they hardly received CPR training, besides, majority of them who indicated that they had not received CPR training expressed that they intended to attend CPR training in the future (84.8%). This result suggests that most nonmedical adults in Muscat have the motivation to receive CPR training. Therefore, it is imperative that awareness campaigns about BLS should be accompanied by training people in the necessary skills for carrying out BLS activities. Moreover, majority of those who indicated that they had received CPR training (29.1%) specified that they had received training from Television-Internet-Media (33.1%), at a course given by the trainers of the Ministry of Health (21.0%), CPR education institutions (12.9%) and at school (12.1%). The proportion of those that had received training is much lower than this proportion in a cross-sectional survey, which was carried out in 5 cities of the Sultanate of Oman (38.9%),^7^ that among an adult sample from a busy street in a city of a western region of Turkey (40.7%),^2^ but comparable with 30.0% which was reported in the UK among Greater London residents,^8^ 35% which was reported in the Japanese general population.^9^ Lower rates of individuals attending BLS training were also reported in Singapore.^10^

### Public knowledge

Majority of the nonmedical adults did not know to find out whether a collapsed person was conscious (69%), or had a pulse (62%), suggesting that they were very unlikely save an unconscious collapsed person, hence a large proportion of unconsciousness due to collapsing in Muscat may have poor outcomes after referral to the hospital. However, substantial number of the participants indicated that they would activate emergency medical service (52.1%) after confirming that the collapsed person was unconscious, and (77.0%) identified the correct number (9999) to call for emergency medical services. This is an encouraging outcome which still needs strengthening in order to increase the number of people who would effectively activate emergency medical services in the face of an emergency. Furthermore, only 32.6% responded correctly on location of chest compressions during CPR. This proportion is far much lower than that among individuals aged ≥18 years on a busy street in a city of a western region of Turkey (52.0%),^2^ but comparable with the proportion of the same among lay-public in the Republic of Slovenia (37.6%).^11^ Additionally, a cross-sectional survey, which was carried out in 5 cities in the Sultanate of Oman, found that 60% of subjects had no appreciation of the need for CPR in a hypothetical scenario in an unconscious child who chocked on a piece of fruit.^7^ However, in this study, a comparable proportion correctly indicated that, upon witnessing an infant choking while playing with a toy but the infant was unable to cry or cough, they would perform back blows and chest compression of five cycles each, then open the mouth and remove the foreign body only when seen (56.6%). In contrast, another comparable proportion incorrectly indicated that, upon observing a friend expressing symptoms of chocking when having a meal, they would give back blows on the victim (57.7%). Thus, a considerable number of nonmedical adults in Muscat do not have prerequisite knowledge for training in first aid skills for helping individuals experiencing chocking and proper interventions in this regard would be cost efficient among this study population. In relation to helping an adult unresponsive victim who had been submerged in fresh water then was just removed from it and was showing signs of spontaneous breathing, but unresponsive, a larger proportion of the participants (93.4%) responded incorrectly on what they would do first in order to help such an adult person. According to the World Health Organization (WHO), drowning is a serious but neglected public health threat that claims the lives of approximately 372, 000 people per year worldwide.^12^

### Association between public knowledge toward BLS and sociodemographic variables

Although the participants were generally poor in knowledge about BLS, this study found that there was an association between some socio-demographic variables and knowledge levels toward BLS specifically, gender, level of education and occupation. For example, male were more knowledgeable than female. Also, Doctoral level holders were more knowledgeable than those other levels of education; whereas those with occupation in Government and Private sectors were more knowledgeable than housewives and students.

The finding of male was more knowledgeable than female is consistent with the findings of some previous study.^13^ Moreover, it is in agreement with the recent findings that the Omani female population is much lagging behind in literate life expectancy than the Omani male population.^14^ Therefore, this finding underscores the need to reduce the gap in BLS knowledge between male and female so that both genders could confidently and skillfully provide BLS services.

Furthermore, just like in this study, several studies have demonstrated that BLS knowledge has been poor among the levels of education, such as primary, secondary, and college.^15 16 17^ Although the doctoral group was associated with better BLS knowledge than the preceding education level groups, their knowledge was still inadequate. Therefore, this outcome suggests that introducing BLS training in the school’s curriculum at all levels would pay great dividends for the prevention of deaths resulting from any emergency life threatening events.

Also, housewives and students demonstrated to have poorer knowledge in BLS than private and government workers is consistent with the findings of other similar studies.^4 18^ Besides, the housewives need not be left out when conducting interventions to improve BLS knowledge among the laypeople of Muscat in Oman. The housewives stay with children at home most of their time and if they do not have adequate skills to provide first aid to fatal accidents involving the kids, then the prevalence of poor outcomes in relation to such type of accidents would increase. Therefore, BLS training for housewives is warranted just as for the other categories of workers.

### Public attitude

It was found that all participants had positive attitude toward BLS. This was an encouraging result, which suggests that many Non-medical people would be willing to attend training in BLS, should there be an initiative to train them.

This finding is consistent with some previous prospective study on senior undergraduate student-teachers enrolled at South African university, which found that the student-teachers surveyed displayed poor knowledge and perceptions but positive attitudes with regards to practice of CPR and BLS, and this suggested that formal CPR training was supposed to be part of the curriculum for teachers.^19^ The BLS training increases laypersons’ confidence and willingness to perform bystander CPR on a stranger. ^20 21^

In this study, despite socio-demographic characteristics being associated with the level of knowledge, they were not associated with the level of attitude. This is not a surprising result considering that all the participants had virtually same level of attitude toward BLS, which is a great factor in making BLS training effective among study population.

### Limitation of the study

Convenience sampling method was used, which is susceptible to selection bias. However, the large sample size might have minimized the possibility of selection bias. Also, cross-sectional design could not establish the causal effect of socio-demographic characteristics on the level of knowledge. Therefore, prospective studies are warranted to confirm the observed associations between socio-demographic characteristics and the level of knowledge. Finally, when performing group comparisons of knowledge and attitude, some groups such as the 48+ years old, the divorced, the primary and doctoral levels of education and the retired groups, had far much smaller subject numbers, which might have compromised the statistical power.

### Implication

This study suggested that there is a huge gap between awareness of BLS and having skills in BLS. Also, the level of knowledge about BLS was very low among the study population but there were still differences in the level of knowledge with respect to the socio-demographic characteristics. In contrast, this study proved that all participants had a positive attitude toward BLS.

Finally, considering the increasing rate of illnesses and injuries that require BLS measures in Oman, policymakers in Oman should advocate mandatory training in BLS for all nonmedical population in the country. Furthermore, incorporating BLS teaching in school curriculum would be helpful for ensuring that many students acquire the necessary knowledge and skills to provide BLS services when needed. Also, learning from and using other people’s experiences in other population as regards BLS training would help increase the knowledge and awareness of BLS, hence increasing the chances that more lives would be saved.

## Data Availability

No data availability

## Acknowledgment

I would like to express my earnest thanks to my research instructor Professor Gu Can for bestowing valuable suggestions and guidance; and for the academic and technical support of Central South University to accomplish this article successfully.

## Funding

no external funding.

## Notes

### Competing Interest Statement

The authors have declared no competing interest.

### Funding Statement

No external funds

## References

1. WIKIPEDIA. Basic life support. 2011.

2. Özbilgin Ş, Akan M, Hancı V, et al. Evaluation of Public Awareness, Knowledge and Attitudes about Cardiopulmonary Resuscitation: Report of İzmir[J]. Turkish journal of anaesthesiology and reanimation, 2015,43(6):396–405.

3. Nadar SK, Mujtaba M, Al-Hadi H, et al. Epidemiology, Outcomes and Coronary Angiography Findings of Patients Following Out-of-Hospital Cardiac Arrest: A single-centre experience from Oman[J]. Sultan Qaboos University medical journal, 2018,18(2):e155-e160.

4. Al-Mohaissen MA. Knowledge and Attitudes Towards Basic Life Support Among Health Students at a Saudi Women’s University[J]. Sultan Qaboos University medical journal, 2017,17(1):e59-e65.

5. Patidar AB, Sharma A. Attitude of school children towards basic life support in Punjab, India. Int J Health Sci Res. 2014;4(5):193–201.

6. Sultan Al-Shaqsi, Ahmed Al-Risi, Ammar Al-Kashmiri. Do Lay People in Oman Know How to Perform Cardiopulmonary Resuscitation?[J]. Oman Med J, 2018,33(2):178–179.

7. Alshaqsi S, Alwahaibi K, Alrisi A. Wasted Potential: Awareness of Basic Cardiopulmonary Resuscitation in the Sultanate of Oman- A Cross-Sectional National Survey [J]. Journal of Emergency Medicine and Intensive Care, 2015,1 (1):105.

8. Donohoe RT, Haefeli K, Moore F. Public perceptions and experiences of myocardial infarction, cardiac arrest and CPR in London[J]. Resuscitation, 2006,71(1):70–79.

9. Kuramoto N, Morimoto T, Kubota Y, et al. Public perception of and willingness to perform bystander CPR in Japan[J]. Resuscitation, 2008,79(3):475–481.

10. Ong MEH, Quah JLJ, Ho AFW, et al. National population based survey on the prevalence of first aid, cardiopulmonary resuscitation and automated external defibrillator skills in Singapore[J]. Resuscitation, 2013,84(11):1633–1636.

11. Rajapakse R, Noč M, Kersnik J. Public knowledge of cardiopulmonary resuscitation in Republic of Slovenia[J]. Wiener klinische Wochenschrift, 2010,122(23):667–672.

12. World Health O. Global report on drowning: preventing a leading killer[M]. Geneva:World Health Organization; 2014.

[13] Krammel M, Schnaubelt S, Weidenauer D, et al. Gender and age-specific aspects of awareness and knowledge in basic life support[J]. PloS one, 2018,13 (6):e0198918-e0198918.

14. M. Mazharul Islam and Md. Hasinur Rahaman Khan. LITERATE LIFE EXPECTANCY AND ITS GENDER DIFFERENTIALS IN OMAN[J]. Journal of Reliability and Statistical Studies, 2019,2(12):21–31.

15. Joseph N, Kumar G, Babu Y, et al. Knowledge of first aid skills among students of a medical college in mangalore city of South India[J]. Annals of medical and health sciences research, 2014,4(2):162–166.

16. Aaberg AMR, Larsen CEB, Rasmussen BS, et al. Basic life support knowledge, self-reported skills and fears in Danish high school students and effect of a single 45-min training session run by junior doctors; a prospective cohort study[J]. Scandinavian Journal of Trauma, Resuscitation and Emergency Medicine, 2014,22(1):24.

17. Almesned A, Almeman A, Alakhtar AM, et al. Basic life support knowledge of healthcare students and professionals in the Qassim University[J]. International journal of health sciences, 2014,8(2):141–150.

18. Ahmad A, Akhter N, Mandal RK, et al. Knowledge of basic life support among the students of Jazan University, Saudi Arabia: Is it adequate to save a life? [J]. Alexandria Journal of Medicine, 2018,54(4):555–559.

19. Ojifinni K, Motara F, Laher AE. Knowledge, Attitudes and Perceptions Regarding Basic Life Support Among Teachers in Training[J]. Cureus, 2019,11(12):e6302-e6302.

20. Pehlivan M, Mercan NC, Çinar İ, et al. The evaluation of laypersons awareness of basic life support at the university in Izmir[J]. Turkish Journal of Emergency Medicine, 2019,19(1):26–29.

21. Abolfotouh MA, Alnasser MA, Berhanu AN, et al. Impact of basic life-support training on the attitudes of health-care workers toward cardiopulmonary resuscitation and defibrillation[J]. BMC Health Services Research, 2017,17(1):674.

